# State-level retrospective analysis of the burden of emergency and operative conditions in India from 1980 to 2023

**DOI:** 10.1101/2025.10.14.25337828

**Authors:** Riley Gerardo, Siddhesh Zadey, Ritika Shetty, Anoushka Arora, Uma Gupta, Shirish Rao, Aiman Perween Afsar, Catherine A. Staton, Joao Ricardo Nickenig Vissoci

**Author notes:** **Corresponding author:** Siddhesh Zadey, BSMS MSc-GH; ASAR Office Address: D2 Sai Heritage, New DP Road, Aundh, Pune, Maharashtra, India 411007.

## Abstract

**Introduction:** Disease burden is a crucial factor in determining healthcare policy and resource allocation. We analyzed the subnational burden of emergency and operative conditions in India from 1980 to 2023.

**Materials and Methods:** We extracted mortality and disability-adjusted life-year (DALY) estimates for 31 Indian states and Union Territories from 1980 to 2023 from the Global Burden of Disease 2023 Study. We used existing expert-consensus-based classifications for defining emergency (30 conditions), operative (57 conditions), and emergency-operative conditions (7 conditions). We investigated rates per 100,000 people and proportions attributable to the above conditions, expressed as percentages of total mortality and DALYs. We conducted a post-hoc uncertainty analysis to calculate the mean and 95% uncertainty interval (UI) for each estimate.

**Results:** In 2023, emergency (mortality rate: 340 deaths per 100,000, 95% UI [282, 416]; DALY rate: 13,732 DALYs per 100,000 people, 95% UI [11,492, 16,277]), operative (mortality rate: 158 deaths per 100,000, 95% UI [123, 196]; DALY rate: 7,284 DALYs per 100,000 people, 95% UI [5,765, 9,018]), and emergency-operative conditions (mortality rate:70 deaths per 100,000, 95% UI [58, 83]; DALY rate: 3,830 DALYs per 100,000 people, 95% UI [3,229, 4,396])accounted for 49.76% (95% UI [43.44%, 58.14%]), 23.12% (95% UI [19.05%, 27.52%]), and 10.31% (95% UI [8.96%, 11.58%]) of all-cause mortality and 40.88% (95% UI [36.91%, 45.40%]), 21.66% (95% UI [18.69%, 24.85%]), and 11.41% (95% UI [10.22%, 12.60%]) of all-cause DALYs, respectively. Telangana, Chhattisgarh, and Uttarakhand had high mortality and DALY rates for all three condition groups. From 1980 to 2023, mortality and DALY rates generally decreased across all conditions, with the largest decreases in emergency conditions. States such as Kerala and Goa did not show significant decreases in their mortality and DALY rates between 1980 and 2023.

**Conclusion:** Despite significant nationwide improvements in healthcare, several states in India have shown a consistent trend over time, reporting a persistently high burden of emergency, operative, and emergency-operative conditions. Given the relatively large share of emergency and operative conditions in all-cause mortality, surgical care should be a high priority in national and state-level cross-sectoral policymaking.

**Highlights:**

- Operative and emergency operative conditions account for 23.12% and 10.31% of all-cause mortality in India, respectively.
- From 1980 to 2023, the burden of operative conditions has decreased at a much slower rate than that of emergency conditions.
- This is the first subnational study of the burden of emergency and operative conditions for India.
- Surgical care should be a high priority in national and state-level cross-sectoral policymaking.

## 1 Introduction

Globally, noncommunicable diseases and injuries account for over 70% of the global mortality burden [1]. A large portion of this burden can be prevented, cured, and managed by adequately strengthening health systems. Previous World Health Organization (WHO) Resolutions [2,3] have codified the centricity of emergency, critical, and operative (ECO) care components within health systems to improve population-level health outcomes and achieve universal health coverage. To accurately measure the magnitude of the problem, prioritize conditions in health policymaking, and allocate resources in an evidence-informed manner, multiple studies have assessed the disease burden of emergency and operative conditions.

Global Burden of Disease (GBD) estimates found that emergency-operative conditions contributed 25 million disability-adjusted life-years (DALYs) and 896,000 deaths globally in 2010 [4]. Another study, which surveyed healthcare practitioners across the globe, found that 25.3% and 28.0% of the global burden of disease is accounted for by deaths and DALYs due to operative conditions [5]. Another study found that essential surgical conditions accounted for 4.7 million deaths and 340 million DALYs in low- and middle-income countries (LMICs) in 2011, and that universal essential surgery coverage would avoid an estimated 1.5 million deaths and 87 million DALYs in LMICs per year [6]. A recent ad hoc analysis using the GBD 2019 Study estimated the global burden of emergency conditions at 367.18 deaths and 13,872.48 DALYs per 100,000 population, compared with the global burden of non-emergency conditions at 368.93 deaths and 17,811.71 DALYs per 100,000 population [7]. They also found a global burden of operative conditions of 233.83 deaths and 8,311.47 DALYs per 100,000 population, as compared to the global burden of non-operative conditions, which was 502.28 deaths and 23,372.68 DALYs per 100,000 population. However, these studies use varying definitions for emergency and operative conditions. For example, Stewart et al. (2014) [4] defined emergency surgical conditions as those most commonly requiring emergency surgery by a general surgeon, excluding causes such as trauma, obstetrics, and orthopedics. In contrast, the DCP3 framework defines essential operative conditions as those primarily or extensively treated by surgical providers, with a large health burden, and that can be effectively treated by surgical procedures/care in a globally cost-effective and feasible manner [6]. Despite differing definitions affecting burden estimates, there is consensus that both emergency and operative care play significant roles in the broader picture of global health, making these areas key to continued research and healthcare reform.

Despite many global studies on the burden of emergency, operative, and emergency-operative conditions, studies on national burdens are limited, and subnational data are lacking. These data would serve as key indicators for national and local governments to develop policies to reduce the burden of emergency and operative conditions. This is particularly necessary for countries with large geographies and complex multi-tier health governance structures, such as India. Additionally, almost all previous global assessments have investigated emergency and operative burdens cross-sectionally, preventing them from commenting on changes over time. Given that India accounts for 17.30% of global mortality and 18.39% of global DALYs [1], we analyzed the burden of emergency, operative, and emergency-operative conditions at the subnational level in India from 1980 to 2023.

## 2 Methods

### 2.1 Data and Definitions

We extracted data from the Global Burden of Disease (GBD) 2023 on deaths, DALYs, and population counts from 1980 (the earliest year available in GBD 2023) through 2023 at the global, national, and subnational (31 states and Union territories) levels [1]. Because there is a cap on the downloadable data from GBD, we combined two extractions: (i) national and subnational data for all years for all ages and both sexes, and (ii) age- and sex-stratified data for India in 2023, spanning 21 age groups and both sexes. Each estimate comprised a posterior mean and a 95% uncertainty interval (UI). We did not have access to the underlying posterior draws at the required stratification, which motivated the post-hoc propagation approach described in Section 2.2.

We calculated aggregated deaths and DALYs for emergency conditions, operative conditions, and emergency-operative conditions using existing disease classification frameworks derived from a Delphi consensus process [7,8–10]. We included 30 emergency conditions that were defined as “a condition that, if not diagnosed and treated within hours to days of onset, often leads to serious physical or mental disability or death”. We included 57 operative conditions and defined them as “any condition that may require the expertise of a surgically trained provider”. Of these conditions, 7 fell under both the emergency and operative categories and were additionally classified as “emergency-operative conditions”. We excluded critical conditions from the analysis because the GBD lacks a globally accepted definition and insufficient data on specific critical illness syndromes, such as sepsis. **Table 1** compares the definitions we used from Wimmer et al. (2025) [7] with those of other recent papers studying the burden of emergency, operative, and emergency-operative conditions. **Figure 1** illustrates the conditions from the GBD 2023 study that were included in each condition group.

**Figure 1:**
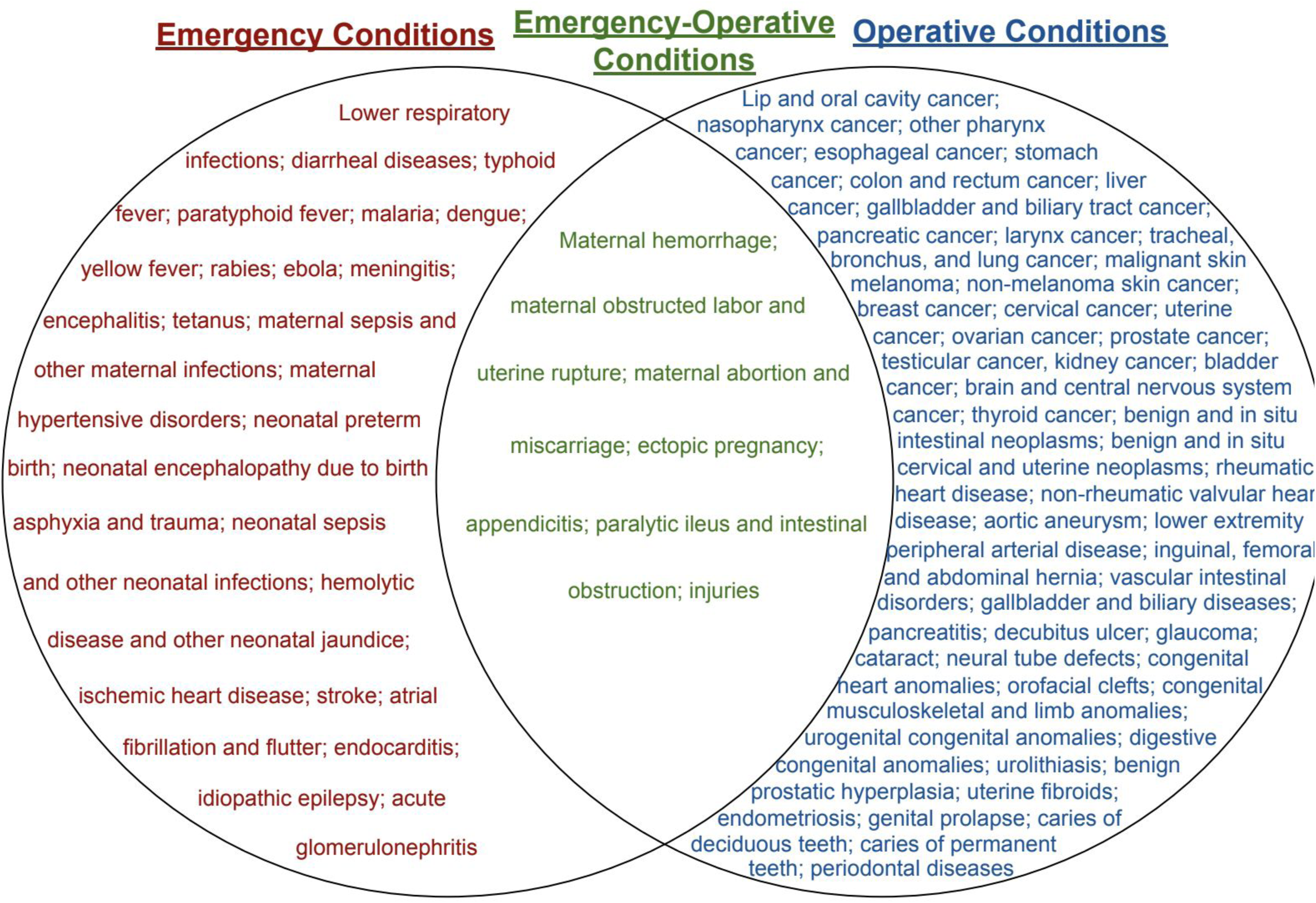
Venn diagram of GBD condition groups based on Wimmer et al. 2025 [7]

**Table 1:** Comparison of Definitions of Emergency, Operative, and Emergency-Operative Conditions.

| Reference | Definition of Emergency Conditions | Definition of Operative Conditions | Definition of Emergency-Operative Conditions |
| --- | --- | --- | --- |
| Wimmer et al., 2025 [7] | “a condition that, if not diagnosed and treated within hours to days of onset, often leads to serious physical or mental disability or death” | “any condition that may require the expertise of a surgically trained provider” | “A condition that was identified as both an emergency condition and an operative condition.” |
| Chang et al., 2016 [10] | Conditions “that, if not diagnosed and treated within hours to days of onset, often lead to serious physical or mental disability or death” |  |  |
| Debas et al., 2015 [6] |  | Conditions “primarily or extensively treated by surgical providers”, with a “large health burden”, and “can be successfully treated by a surgical procedure and other surgical care that is cost-effective and feasible to promote globally”* |  |
| Stewart et al., 2014 [4] |  |  | Conditions that are most commonly an indication for emergency surgery by a general surgeon,<br>excluding trauma,<br>burns, obstetrics,<br>neurosurgery,<br>orthopedics,<br>complex congenital<br>surgery, and<br>cardiothoracic<br>surgery |

\* The definition from Debas et al. is for essential surgical conditions, not surgical conditions as a whole.

### 2.2 Primary Analysis

We grouped data by year, location, measures (death or DALYs), age groups (5-year bins from <5 to 95+ years), sexes (males and females), and condition groups (emergency, operative, or emergency-operative). We estimated the following outcomes: the total count, rate per 100,000 people, and % of all-cause deaths/DALYs **(Table 2)**. For each outcome, we report the point estimate (the mean-based estimate or the mean of the draw distribution for derived quantities) together with a 95% UI defined by the 2.5^th^ and 97.5^th^ percentiles of the draws.

**Table 2:** Calculations for outcomes.

| Outcome | Equation |
| --- | --- |
| Rate per 100,000 people | $Rate_{y,l,m,g} = \frac{total\ \#_{y,m,l,g}}{population_{y,l}} \times 100,000$ |
| % of all-cause deaths or DALYs | $\% of\ all\ cause = \frac{total\ \#_{y,m,l,g}}{total\ \# of\ all - cause_{y,m,l}} \times 100$ |

y=year, l=location, m=measure (Deaths or DALYs), and g=condition group

The GBD 2023 did not provide posterior draws for the stratifications used in this analysis, which prevents us from estimating uncertainty intervals for the outcomes of interest by simply aggregating GBD estimates. Therefore, we conducted a post hoc Monte Carlo propagation that reconstructed an approximate distribution for each reported estimate from its mean and 95% UIs, drew pseudo-samples, and passed them through the analytic pipeline. Point estimates came from the GBD means; intervals came from the 2.5th and 97.5th percentiles of the pseudo-draws. These intervals approximate the propagation of GBD uncertainty but do not reproduce the full GBD posterior covariance structure.

For each estimate, we fit a three-parameter shifted lognormal distribution so that the fitted distribution reproduced the reported mean and both 95% interval bounds exactly. This approach works well for right-skewed positive count distributions, whose mean is typically below the midpoint of its interval. For estimates that did not follow a lognormal distribution well, we used the following alternatives: truncated normal for near-symmetric estimates, a quantile-matched lognormal for extreme skew, and a degenerate handler for zero or malformed values. Every reconstruction was a monotone transformation of the underlying standard normal draw, which preserved the sign of correlations when the draws were correlated.

Since aggregate quantities (e.g., national totals, condition-group totals) are sums of many individual estimates, the width of a sum depends on the correlation among its components. Rather than assuming this correlation, we calibrated it to GBD’s published UIs, available at both the component and aggregate levels. We computed variances on the natural (count) scale, where aggregation is additive. Based on the available data, we calibrated two correlations by this method: a cross-state correlation, from each condition’s state-level intervals versus the corresponding India national interval, and a within-state, cross-condition correlation, from each state’s condition-level intervals versus that state’s all-cause interval. We pooled calibrated values (median) across strata, with dispersion providing a sensitivity range. Applied to the observed data, the cross-state correlation was moderate to strong (approximately 0.50 to 0.85).

To ensure that state estimates sum to the national estimate for point estimates and intervals, we treated the most granular data cell (state/UT × year × condition, at all-ages/both-sexes) as the only primitive random quantity and derived every aggregate by summation within each draw. We obtained national totals by summing draws across the 31 subnational regions, and condition-group totals by summing the individual conditions. Because counts are additive, the summed component means should exactly equal the published aggregate means. The calibrated correlations ensure the summed intervals reproduce the published aggregate widths. Within each draw, the component-level shocks followed a crossed random-effects structure: a condition factor shared across states (inducing the calibrated cross-state correlation) and a state factor shared across individual conditions (inducing the within-state correlation), with independent residual variation. This construction makes states add to the nation within a draw by design.

We used age- and sex-stratified data, only for India in 2023. Therefore, we did not make it subnationally consistent. We report it as a descriptive decomposition of the national total. To ensure the age-sex components added up to the nationally consistent total, we normalized each draw of the age-sex estimates to sum to one, and multiplied these shares by the corresponding national total draw. The resulting age-sex burdens sum to the national total exactly in every draw. Each age-sex mean equals the product of the GBD age-sex share and the consistent national mean.

We calculated the share of all-cause burden attributable to a condition group within each draw, where all-cause total deaths/DALYs were taken directly from GBD. The denominator was constructed so that the numerator is always a subset of it: the all-cause draw was decomposed into the condition group plus a remainder term (with mean equal to GBD all-cause minus the group means), and the remainder was drawn and added to the group draw.

This preserved the correct positive dependence between numerator and denominator and kept the proportion within the 0 to 100 range. The reported intervals are the 2.5^th^ and 97.5^th^ percentiles of the per-draw proportions.

We performed the analysis in R version 4.3.1. We used Claude Cowork Opus 4.8 for statistical simulations and code validation. All data and code used for this study are available at the Zenodo repository [11].

## 3 Results

### 3.1 Deaths

#### 3.1.1 Emergency Conditions

We found that 4,908,642 deaths (95% UI [4,066,654, 6,004,923]) in India in 2023 were attributed to emergency conditions, accounting for 49.76% (95% UI [43.44%, 58.14%]) of all-cause mortality. In 2023, India had a mortality rate of 340 deaths per 100,000 people (95% UI [282, 416]) due to emergency conditions, just below the global rate of 345 deaths per 100,000 (95% UI [306, 389]). The national mortality counts in India due to emergency conditions were highest in those aged 70-74 years old, and were generally higher in males than in females, except for those >74 years old (**Figure 2A)**. Overall, India’s national mortality rate decreased from 1980 to 2023 **(Figure 2B)**. In contrast, the mortality rates in Goa and Kerala increased over time **(Figure 2C)**. Mortality rates in several states, including Telangana, Andhra Pradesh, and Chhattisgarh, did not decrease substantially from 2003 to 2023. Among all locations, Uttar Pradesh, Madhya Pradesh, Odisha, and Bihar showed large overall decreases in mortality rates between 1980 and 2023. In 2023, 12 states in India had mortality rates above the national average, whereas 18 states had rates below it **(Figure 2D)**. In 2023, the states with high mortality rates were Telangana, Goa, and Andhra Pradesh. In comparison, Mizoram, Arunachal Pradesh, and Meghalaya had low mortality rates due to emergency conditions in 2023.

**Figure 2:**
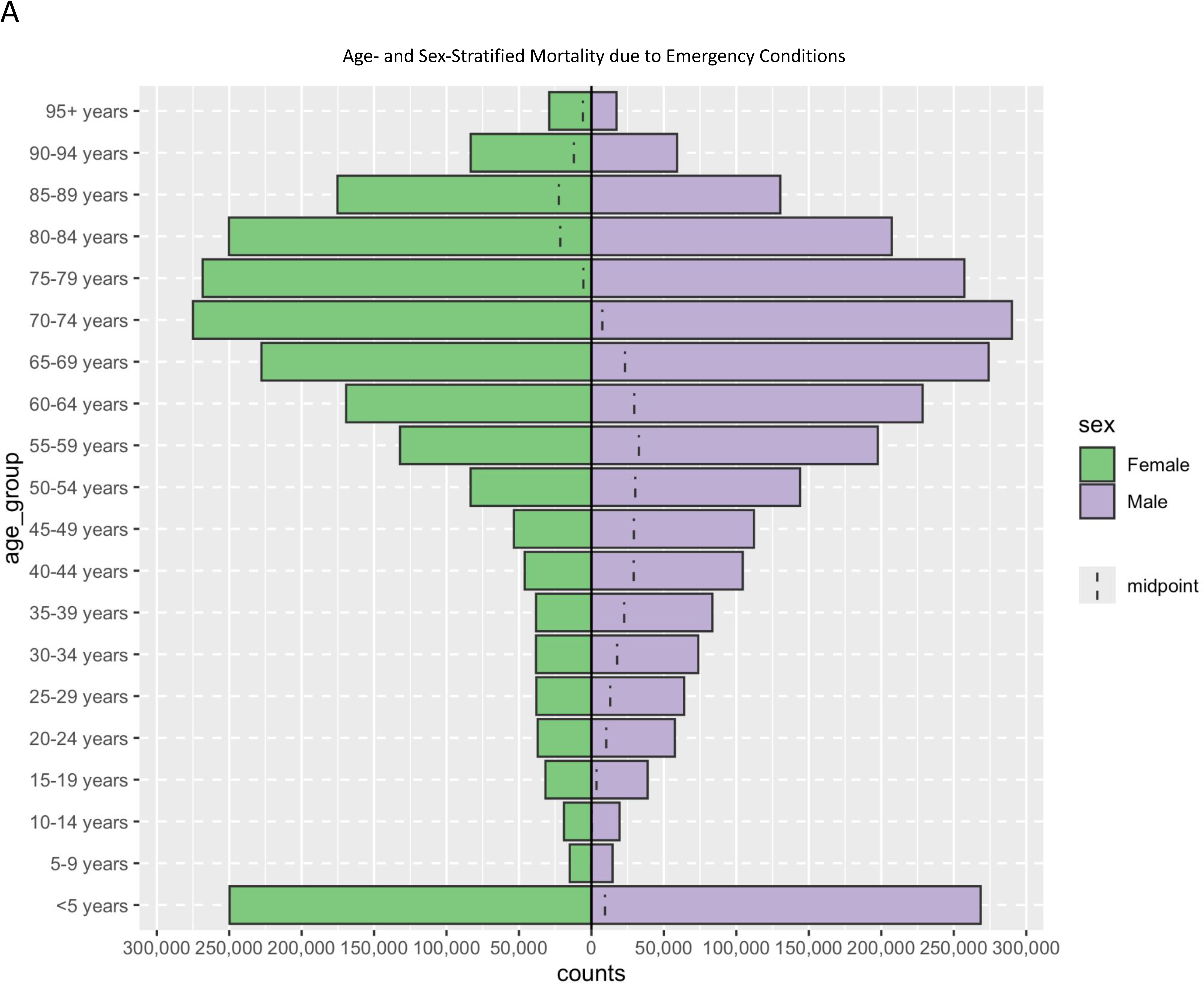
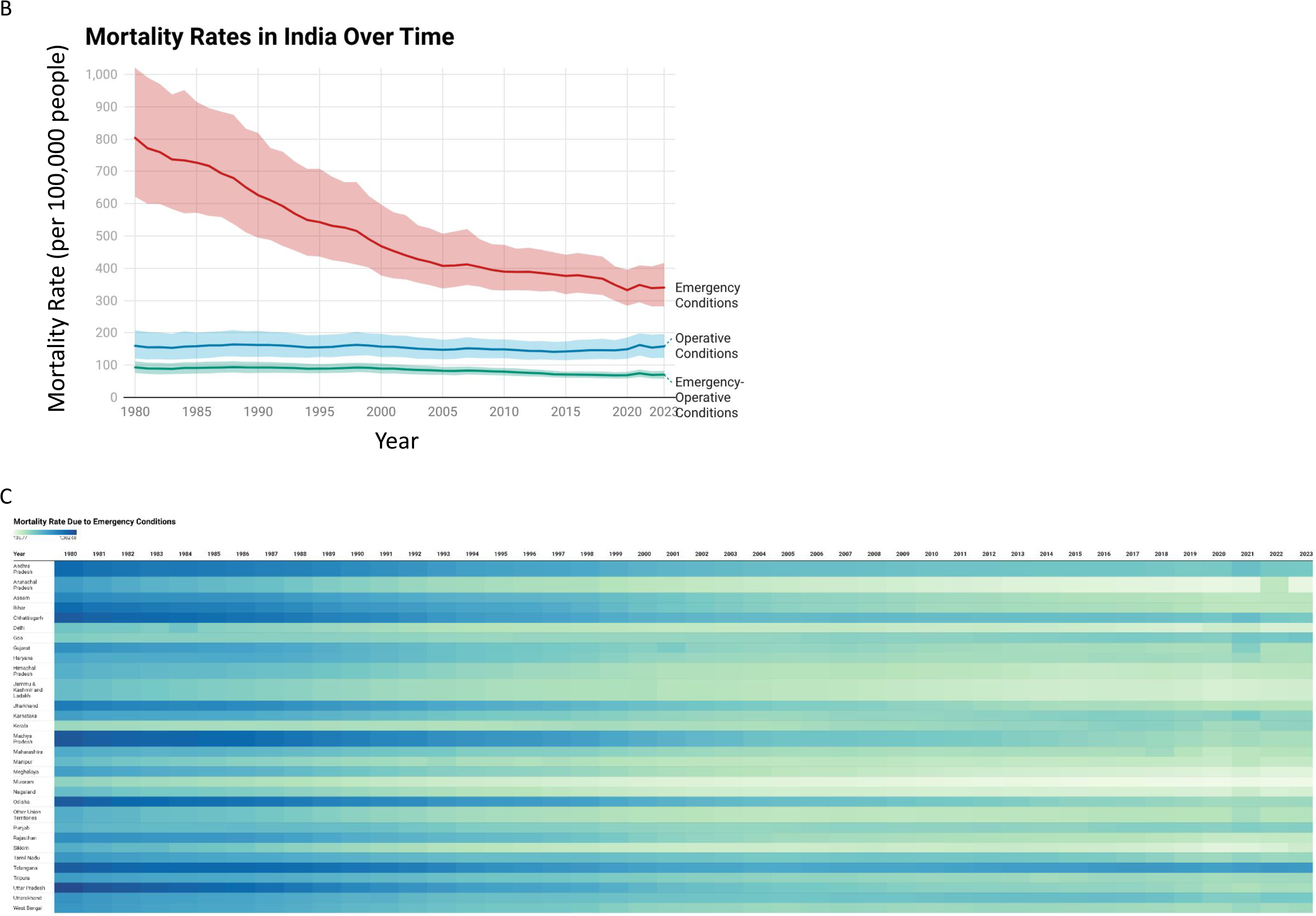
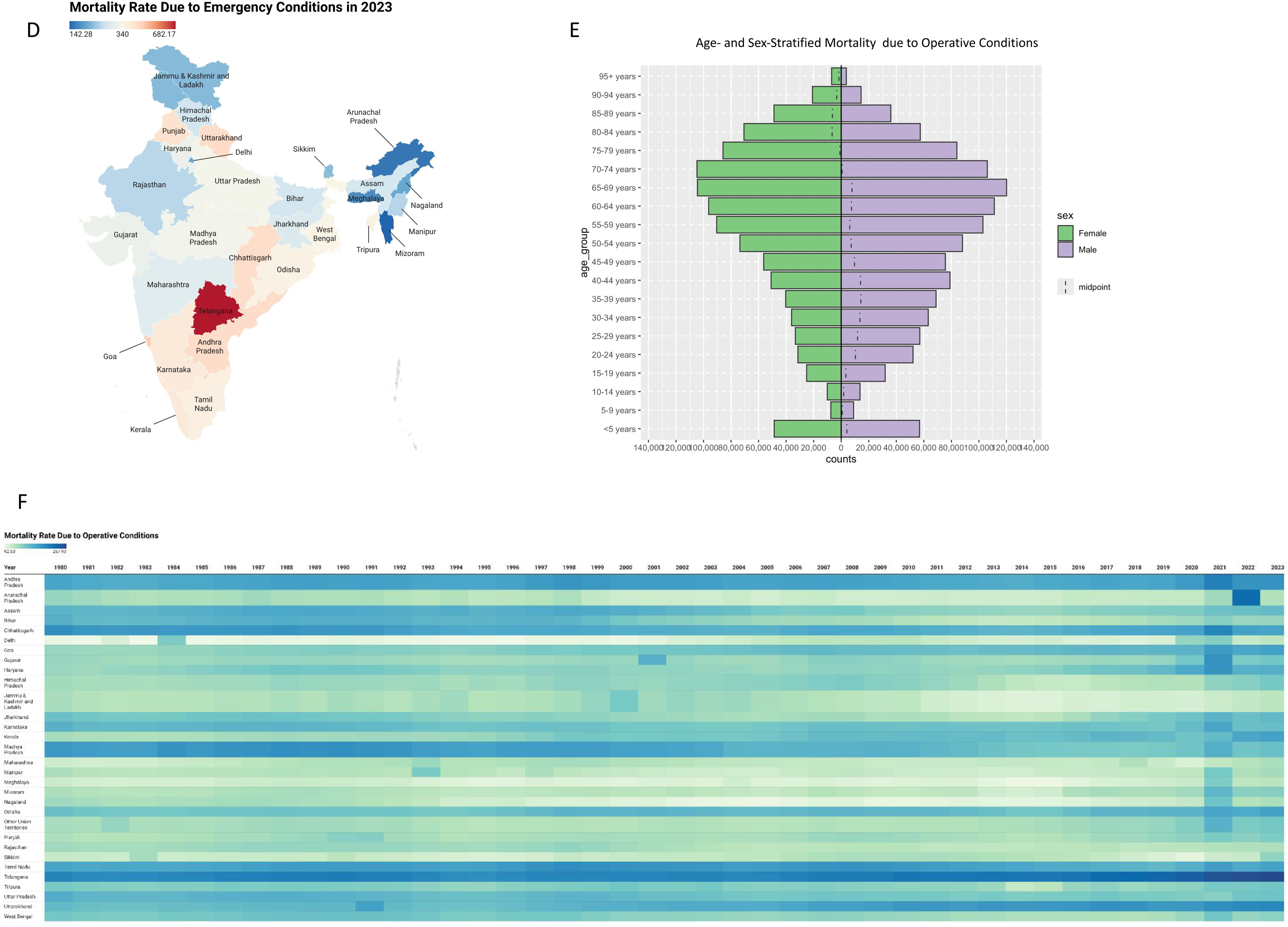
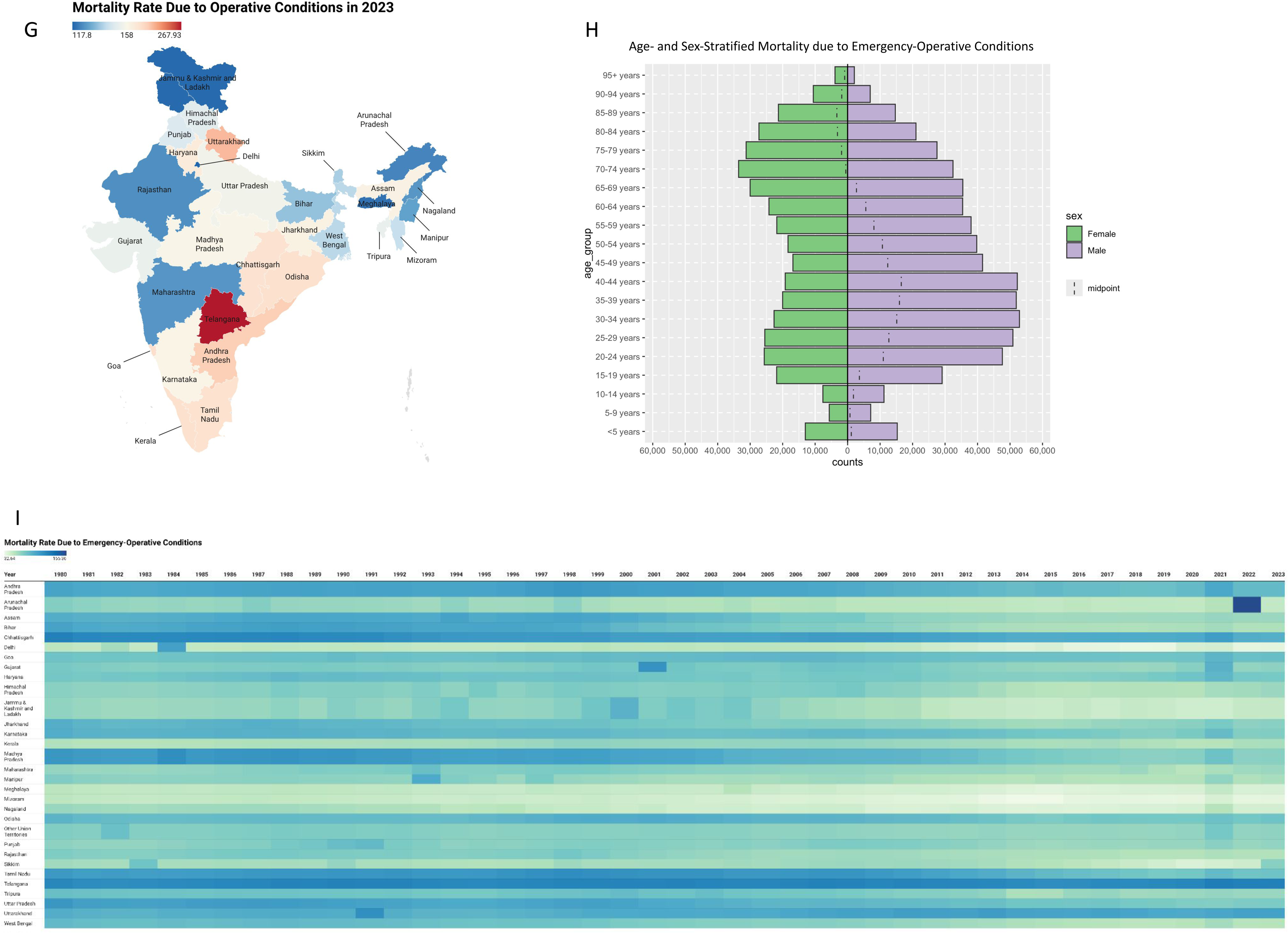
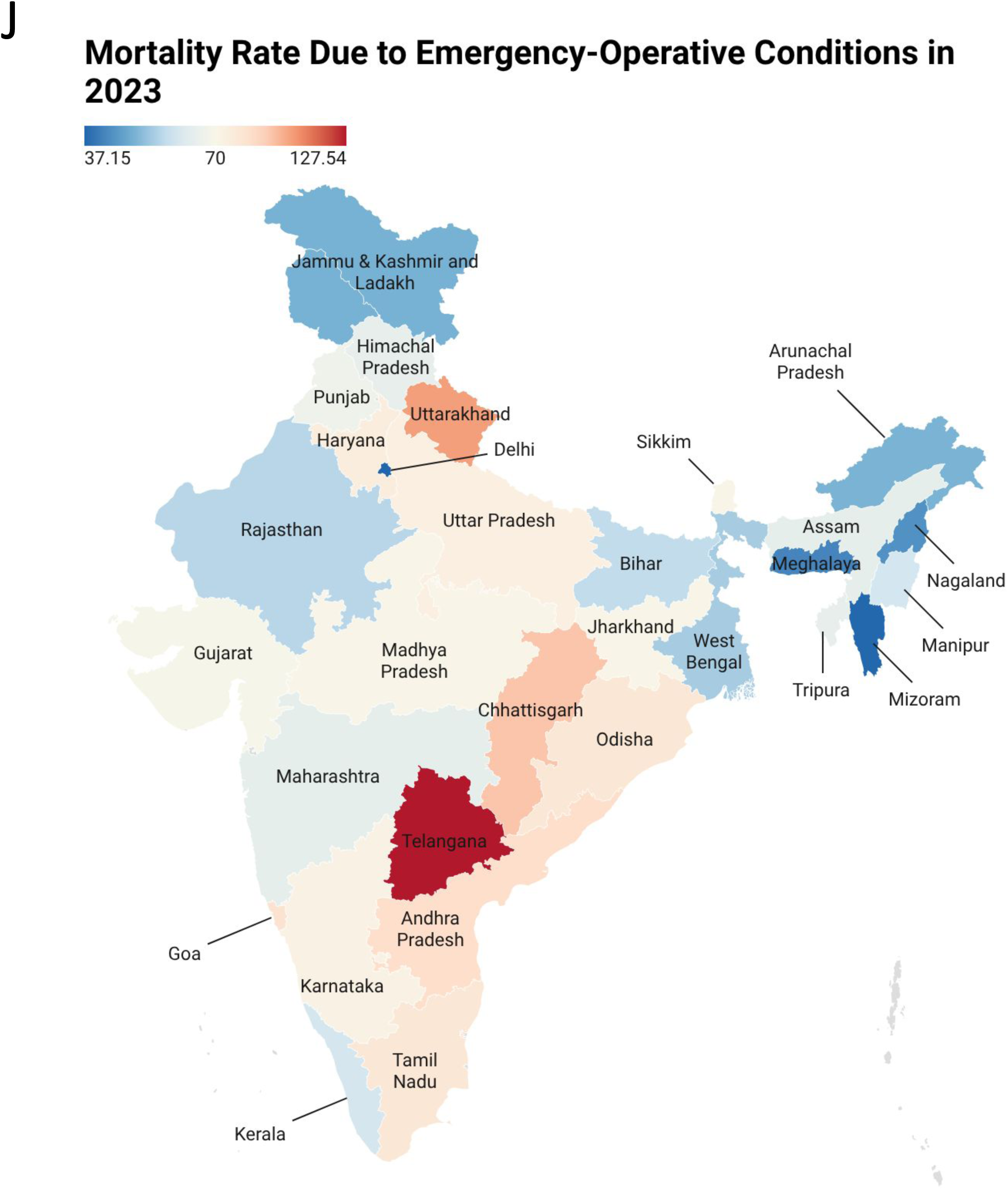
A) Age- and Sex-Stratified Mortality Counts due to Emergency Conditions in India in 2023. B) National Mortality Rates in India over time. C) Heatmap of mortality rates due to emergency conditions. D) Map of mortality rates due to emergency conditions across India in 2023. E) Age- and Sex-Stratified Mortality Counts due to Operative Conditions in India in 2023. F) Heatmap of mortality rates due to operative conditions. G) Map of mortality rates due to operative conditions across India in 2023. H) Age- and Sex-Stratified Mortality Counts due to Emergency-Operative Conditions in India in 2023. I) Heatmap of mortality rates due to emergency-operative conditions. J) Map of mortality rates due to emergency-operative conditions across India in 2023.

#### 3.1.2 Operative Conditions

We found a total of 2,281,829 deaths (95% UI [1,776,09, 2,830,748]) from operative conditions in India in 2023, accounting for 23.12% of all-cause mortality (95% UI [19.05%, 27.52%]). India had a national mortality rate of 158 deaths per 100,000 (95% UI [123, 196]) in 2023, which is below the global rate of 201 deaths per 100,000 (95% UI [180, 225]) in 2023. The national mortality counts in India due to operative conditions were highest in those aged 65-69 years and, as seen with emergency mortality counts, were generally higher in males than in females, except for those >74 years old (**Figure 2E)**. Overall, the mortality rate due to operative conditions in India decreased from 1980 to 2023, although it started much lower and declined less than the mortality rate due to emergency conditions **(Figure 2B)**. In contrast, 20 states, including Telangana, Goa, and Kerala, experienced an overall increase in mortality rates from 1980 to 2023 **(Figure 2F)**. Furthermore, several states, such as Tamil Nadu, Assam, and West Bengal, did not experience substantial decreases in mortality rates from 1980 to 2023. Bihar, Madhya Pradesh, and Uttar Pradesh showed the largest overall decreases in mortality over time. In 2023, 13 states had mortality rates above the national average, and 17 had rates below it **(Figure 2G)**. Telangana, Uttarakhand, and Andhra Pradesh had high mortality rates in 2023. In comparison, Jammu and Kashmir and Ladakh, Delhi, and Meghalaya had low mortality rates in 2023.

#### 3.1.3 Emergency-Operative Conditions

We found 1,016,437 deaths (95% UI [833,631, 1,192,737]) due to emergency-operative conditions in India in 2023, accounting for 10.31% of all-cause mortality (95% UI [8.96%, 11.58%]). For emergency-operative conditions, India had a national mortality rate of 70 deaths per 100,000 (95% UI [58, 83]), slightly above the global rate of 65 deaths per 100,000 (95% UI [59, 71]) in 2023. The national mortality rate in India due to emergency-operative conditions was highest in those aged 30-34 years old, and was generally higher in males than in females, except for those aged 70+ years old (**Figure 2H**). Overall, the mortality rate due to emergency-operative conditions in India decreased from 1980 to 2023, although it started much lower and decreased much less than the mortality rate due to emergency conditions in India **(Figure 2B)**. In contrast, the mortality rate in Kerala, Telangana, Goa, Sikkim, and Uttarakhand increased between 1980 and 2023 **(Figure 2I)**.

Some states reached their peak mortality rates after 1980. For example, Gujarat peaked in 2001. Several states also had periods when their mortality rates did not decrease much. For example, the mortality rate in several states, including Jharkhand, Jammu and Kashmir, and Ladakh, did not decrease much from 2013 to 2023. Bihar, Madhya Pradesh, and Uttar Pradesh experienced large overall decreases in mortality rates between 1980 and 2023. In 2023, 13 states had mortality rates above the national average, and 17 had rates below it **(Figure 2J)**. Telangana, Uttarakhand, and Chhattisgarh had high mortality rates in 2023, while Mizoram, Delhi, and Meghalaya had low mortality rates.

### 3.2 DALYs

#### 3.2.1. Emergency Conditions

We found a total of 198,210,604 DALYs (95% UI [165,867,397, 234,936,279]) in India in 2023 attributable to emergency conditions, accounting for 40.88% of all-cause DALYs (95% UI [36.91%, 45.40%]) lost in India that year. In 2023, India had a national DALY rate of 13,732 DALYs per 100,000 people (95% UI [11,492, 16,277]), slightly above the global rate of 13,670 DALYs per 100,000 people (95% UI [11,857, 15,737]). In India, national DALY counts from emergency conditions were highest among those aged <5 years and higher in males than in females, except among those >74 years old (**Figure 3A**). Overall, India’s national DALY rate decreased over time **(Figure 3B)**. More specifically, the DALY rates in Kerala and Goa decreased only slightly over time **(Figure 3C)**. In contrast, Madhya Pradesh, Bihar, and Uttar Pradesh had large decreases in DALY rates between 1990 and 2023. In 2023, 20 states had DALY rates below the national average, while only 10 were above it **(Figure 3D)**. Telangana, Chhattisgarh, and Uttarakhand had high DALY rates in India in 2023. In contrast, Mizoram, Arunachal Pradesh, and Delhi had low DALY rates in 2023.

**Figure 3:**
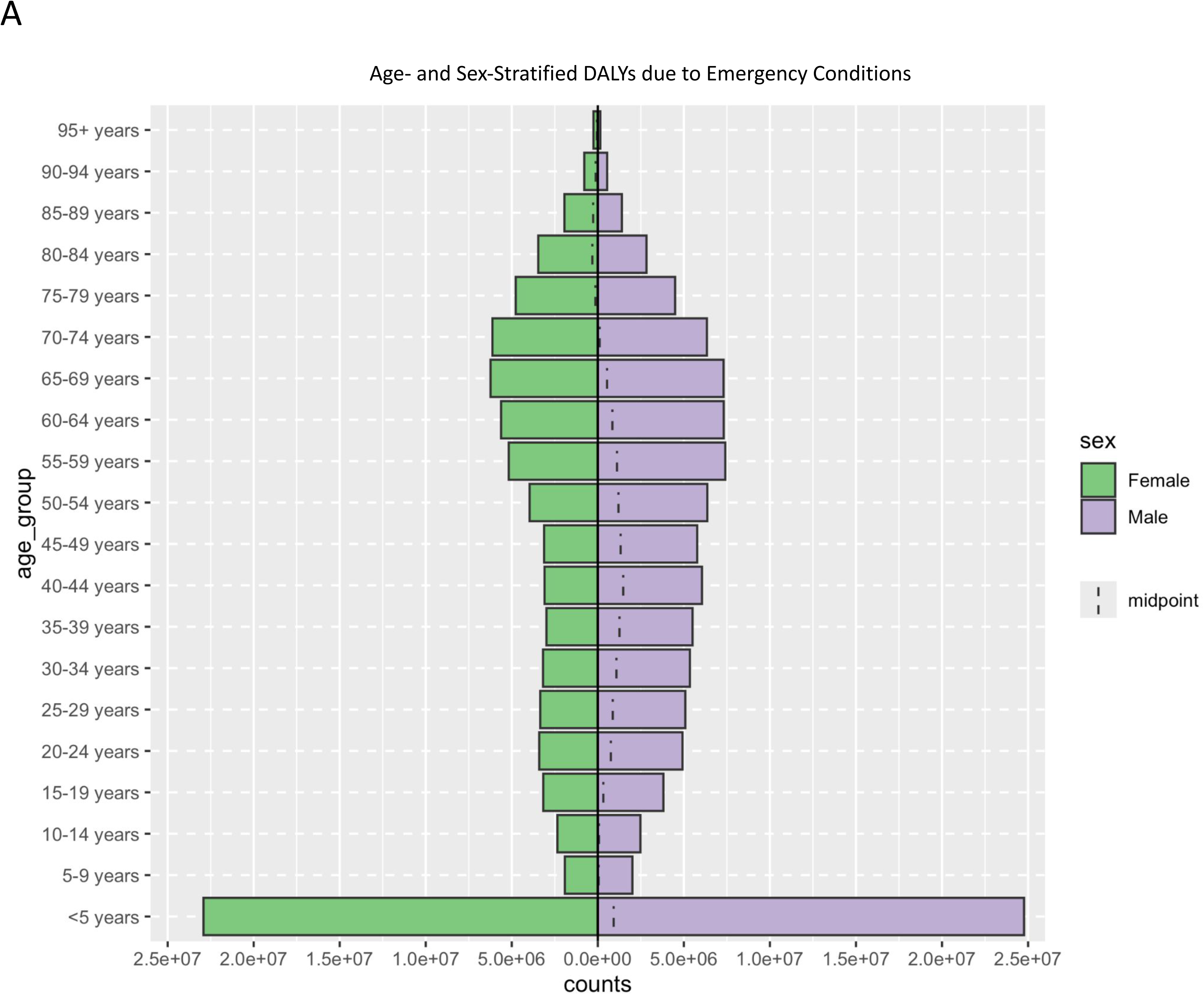
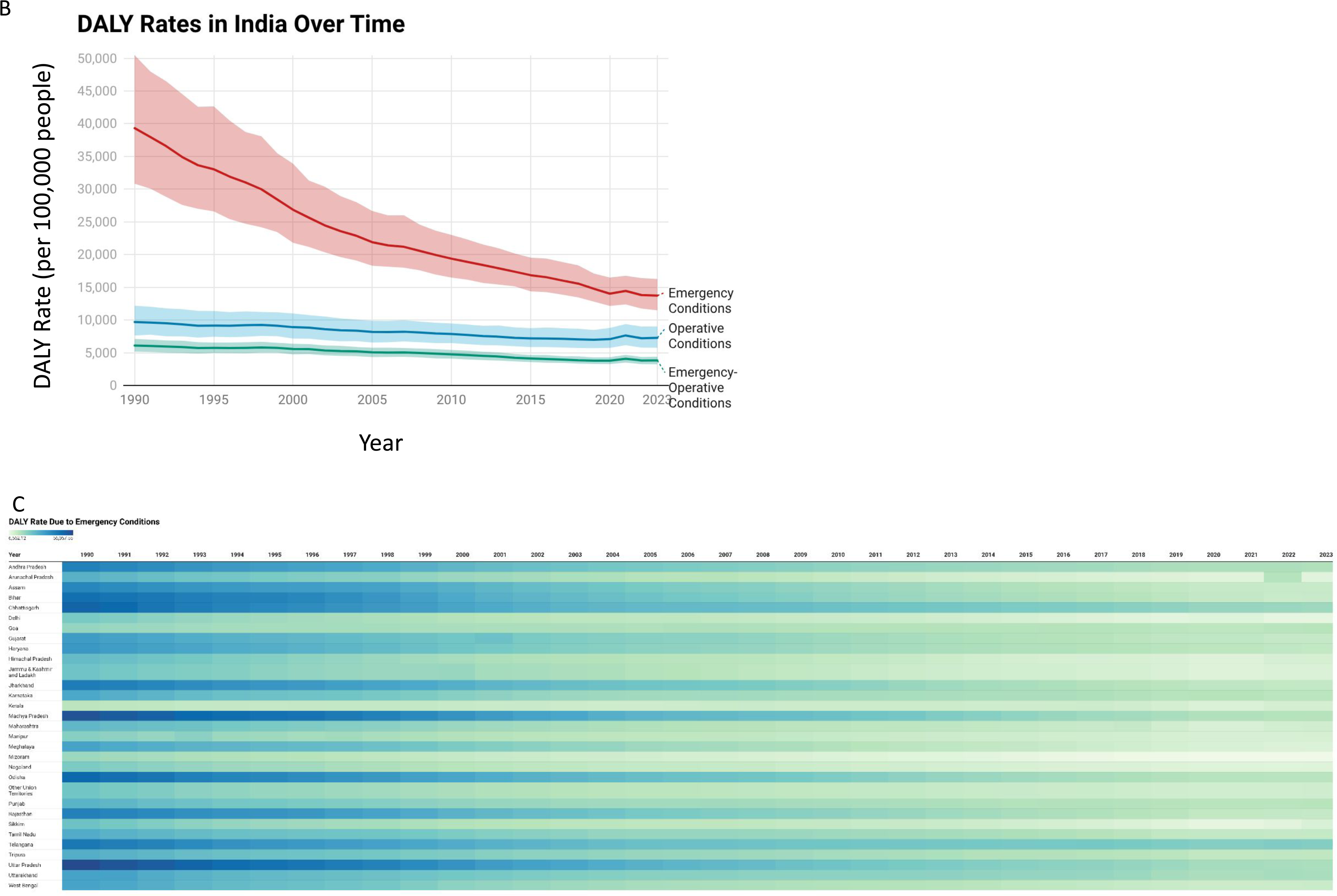
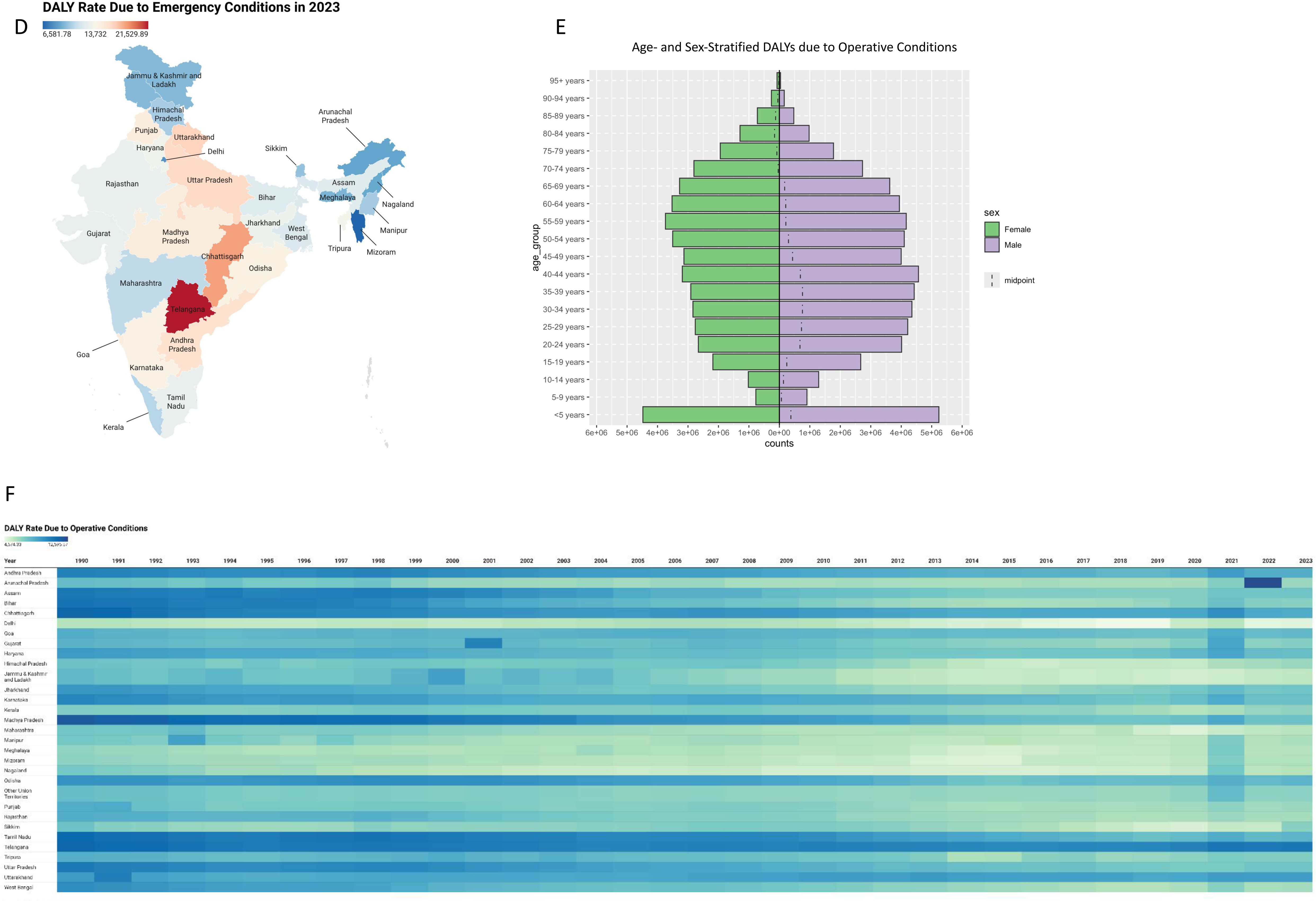
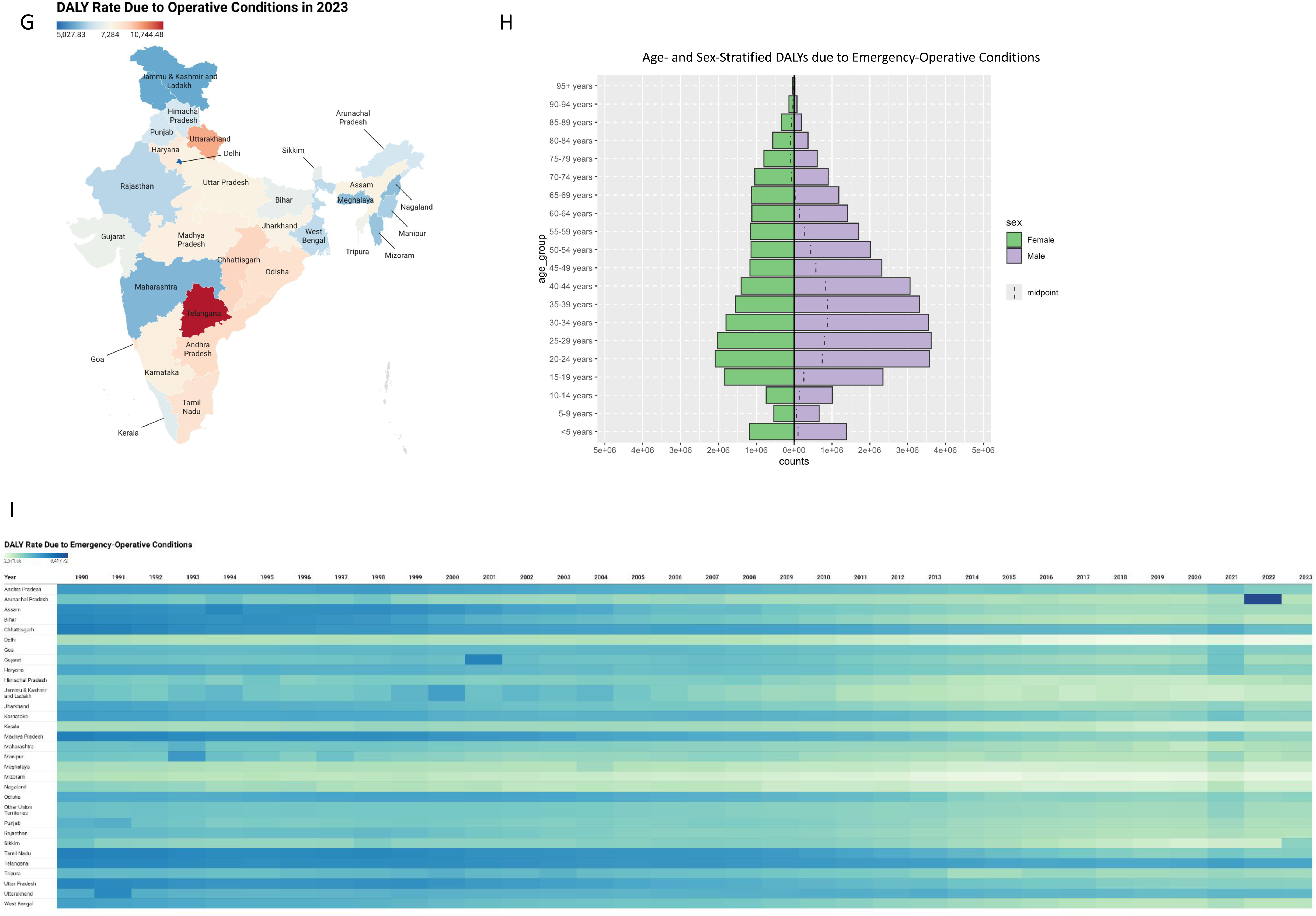
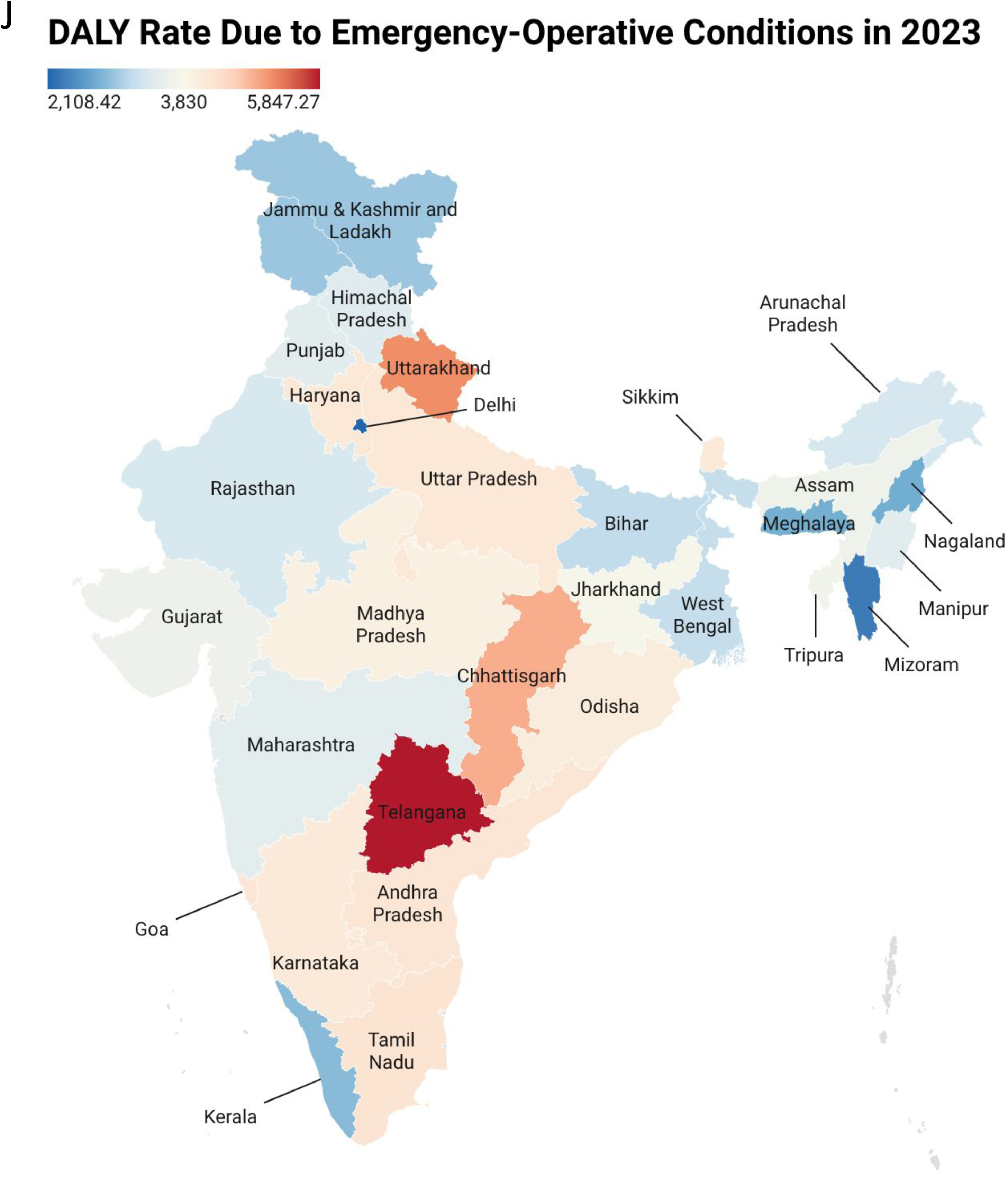
A) Age- and Sex-Stratified DALY Counts due to Emergency Conditions in India in 2023. B) National DALY Rates in India over time. C) Heatmap of DALY rates due to emergency conditions. D) Map of DALY rates due to emergency conditions across India in 2023. E) Age- and Sex-Stratified DALY Counts due to Operative Conditions in India in 2023. F) Heatmap of DALY rates due to operative conditions. G) Map of DALY rates due to operative conditions across India in 2023. H) Age- and Sex-Stratified DALY Counts due to Emergency-Operative Conditions in India in 2023. I) Heatmap of DALY rates due to emergency-operative conditions. J) Map of DALY rates due to emergency-operative conditions across India in 2023.

#### 3.2.2 Operative Conditions

We found a total of 105,130,980 DALYs (95% UI [83,204,611, 130,169,833]) due to operative conditions in India in 2023, which accounted for 21.66% of all-cause DALYs (95% UI [18.69%, 24.85%]). For operative conditions, India had a national DALY rate of 7,284 DALYs per 100,000 people (95% UI [5,765, 9,018]) in 2023, which was below the global DALY rate of 8,316 per 100,000 people (95% UI [7,266, 9,600]) in 2023. The national DALY rate in India due to operative conditions was highest among those aged <5 years and was generally higher in males than in females, except for those >70 years old (**Figure 2E)**. Overall, the DALY rate due to operative conditions in India decreased from 1990 to 2023; however, it did not decrease as much or start as high as the DALY rate due to emergency conditions in India **(Figure 3B)**. Notably, the DALY rate in Uttarakhand increased slightly between 1990 and 2023, whereas those of Kerala and Goa decreased only slightly over the same period **(Figure 3F)**. Several states, such as Odisha, Haryana, Jharkhand, and Madhya Pradesh, did not experience a substantial decrease in DALY rate between 2013 and 2023. A few states also reached peak DALY rates after 1990, including Gujarat (2001) and Jammu and Kashmir and Ladakh (2000). Madhya Pradesh and Bihar had large overall decreases in DALY rates over time. In 2023, 17 states had a DALY rate below the national average **(Figure 3G)**. Of these states, Uttarakhand, Chhattisgarh, and Telangana had high DALY rates, while Delhi, Jammu and Kashmir and Ladakh, and Maharashtra had low DALY rates.

#### 3.2.3 Emergency-Operative Conditions

We found a total of 55,282,853 DALYs (95% UI [46,609,096, 63,451,249]) due to emergency-operative conditions in India in 2023, accounting for 11.41% of all-cause DALYs (95% UI [10.22%, 12.60%]). India’s national DALY rate of 3,830 DALYs per 100,000 people (95% UI [3,229, 4,396]) in 2023 was slightly below the global rate of 4,092 DALYs per 100,000 people (95% UI [3,612, 4,603]). The national DALY rate in India due to emergency-operative conditions was highest in those aged 25-29 years old, and was higher in males than in females, except for those >70 years old (**Figure 3H)**. Overall, the national DALY rate due to emergency-operative conditions in India decreased between 1990 and 2023; however, it decreased by a smaller margin than the DALY rate due to emergency conditions and started much lower **(Figure 3B**). Within India, the DALY rates in Meghalaya, Kerala, Sikkim, and Uttarakhand decreased the least over time **(Figure 3I)**. Jammu and Kashmir and Ladakh peaked in 2000, and Gujarat peaked in 2001. Bihar, Madhya Pradesh, Assam, and Tamil Nadu showed large overall decreases in DALY rates over time. In 2023, 18 states in India had a DALY rate below the national average **(Figure 3J)**. Uttarakhand, Telangana, and Chhattisgarh had high DALY rates in 2023, while Mizoram, Delhi, and Nagaland had low DALY rates.

## 4 Discussion

### 4.1 Summary of Findings

We showed that emergency conditions (49.76% of all-cause mortality; 40.88% of all-cause DALYs) and operative conditions (23.12% of all-cause mortality; 21.66% of all-cause DALYs) account for a large share of deaths and DALYs in India. The death counts due to emergency and operative conditions were generally highest in those aged 65-74 years old, while the DALY counts for these condition groups were generally highest in those <5 years old. The death and DALY counts for emergency-operative conditions were generally highest for those around 25-34 years old. The death and DALY counts for all condition groups were typically higher in males than in females, except in populations aged 70+ years. Overall, mortality and DALY rates for emergency, operative, and emergency-operative conditions decreased in India from 1980 to 2023. However, some exceptions existed. Mortality rates from emergency conditions increased over time in both Goa and Kerala **(Figure 2C)**. Mortality rates from operative conditions across 20 states, including Telangana, Goa, and Kerala, increased over time **(Figure 2F)**. Mortality from emergency-operative conditions in Kerala, Telangana, Goa, Sikkim, and Uttarakhand increased over time **(Figure 2I)**. Our results suggest that the burden of emergency and operative conditions, and improvements in that burden, have not been equal across all Indian states. Telangana, Chhattisgarh, and Uttarakhand had some of the highest mortality and DALY rates across all 3 condition groups **(Figure 4)**. Delhi, Meghalaya, and Mizoram had some of the lowest mortality and DALY rates across all 3 condition groups **(Figure 4)**.

**Figure 4:**
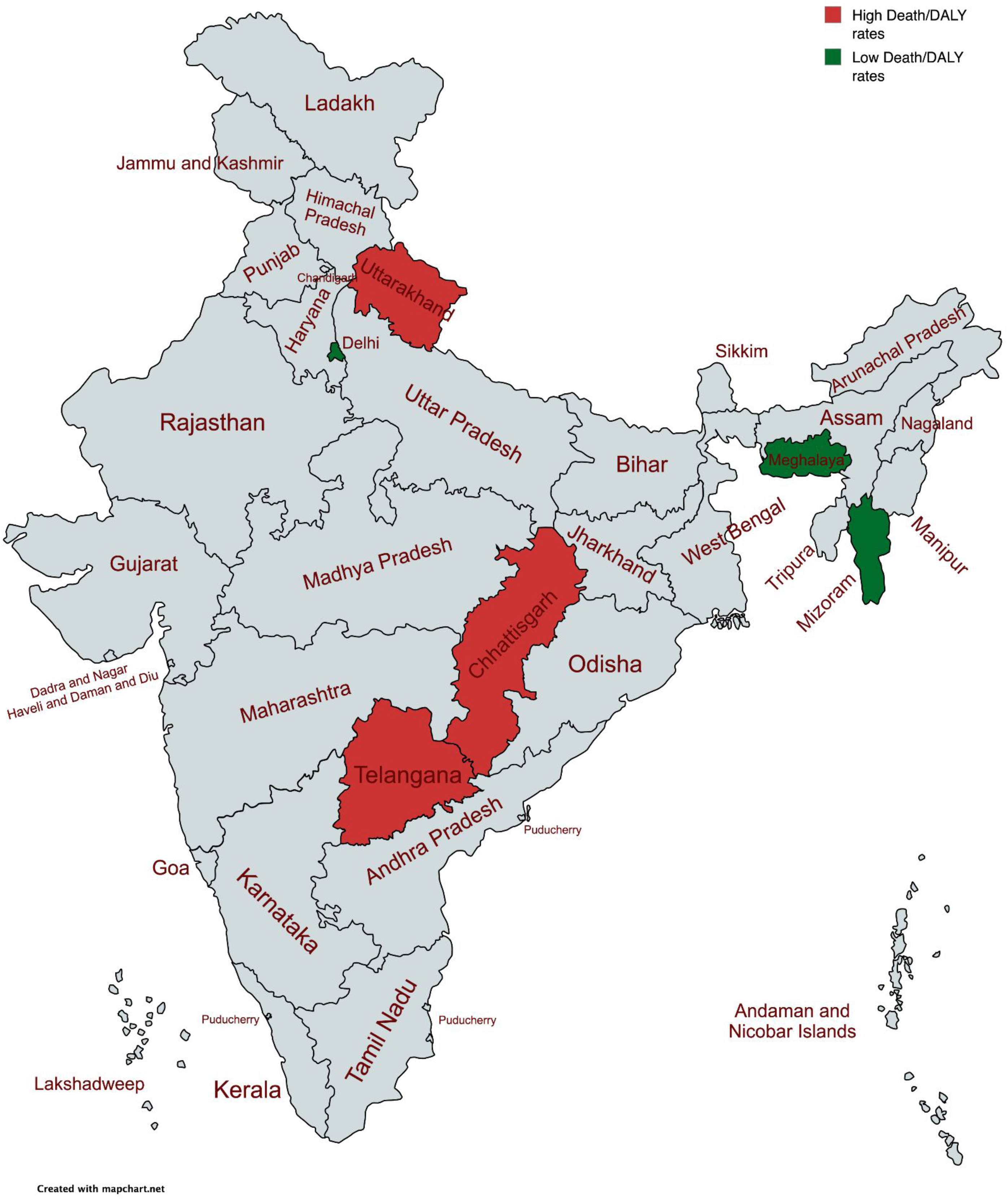
Map of states with high mortality and DALY rates for all three condition groups vs states with low mortality and DALY rates for all three condition groups.

### 4.2 Policy Implications

Several studies have found that emergency and operative care in India lacks accessibility and quality [12,13,14,15]. A systematic review of Indian policies and programmatic guidelines from 1946 to 2017 revealed low prioritization of surgical, obstetric, trauma, and anesthesia care, with a notable decline in policy prioritization after 2010 [16]. Additionally, several studies have found that India falls well below the desired threshold for the six surgical care indicators set out by the Lancet Commission on Global Surgery (LCoGS), and that these indicators vary across Indian states and Union Territories [17,18]. These studies, along with our findings of a high burden of emergency and operative conditions, emphasize the need for improved policy on emergency and operative care in India.

There is a clear need to scale up the healthcare system to address emergency and operative care needs in India, with particular emphasis on states with a high burden of emergency and operative conditions and/or poor performance on the LCoGS indicators. Policy and resource allocation should target states with high mortality rates and concerning long-term trends. States with high mortality rates due to emergency conditions, such as Telangana, Goa, and Andhra Pradesh, should be prioritized for investment to strengthen emergency care, including pre-hospital emergency medical services, standardized trauma protocols, improved resource allocation, and the emergency care workforce.

For operative conditions, Telangana, Uttarakhand, and Andhra Pradesh had high mortality rates in 2023, indicating failure in either perioperative, surgical, or postoperative care. Policies in these states must prioritize expanding the surgical workforce, potentially through incentivized specialist postings in underserved areas. Moreover, states must ensure adequate resources and budget allocation to strengthen infrastructure. Goa and Kerala, which have seen increases in mortality rates for emergency conditions despite a decline in national mortality rates from 1980 to 2023, need a thorough audit of the systemic challenges driving this upward trend. Similarly, states such as Telangana, Goa, and Kerala saw increases in mortality rates among operative conditions. These states should prioritize funding for emergency medicine training/education, mapping emergency centers, and assessing access to these centers.

Furthermore, the Lancet Commission on Global Surgery (LCoGS) recommends that LMICs develop a National Surgical, Obstetric, and Anesthesia Plan (NSOAP) to ensure equitable access to care [19]. While several countries in Africa and South Asia have developed NSOAPs, India has not yet prepared one. Several elements of policymaking, planning, and service delivery in India differ across states. However, State Governments often follow the Central Government’s lead to align state-level efforts with the national agenda. Regardless, states with technical and implementation capacity, particularly those facing relatively high burdens of emergency, operative, and emergency operative conditions, could benefit from urgent state-level policy action.

Our study examined population-level health outcomes influenced by factors within and outside the healthcare system. Thus, we cannot assume that our findings of a high burden of emergency and operative conditions in some states are solely attributable to healthcare system failures, or that new healthcare policies alone would reduce the burden in these states. Future studies could explore how specific policies and socioeconomic trends have driven the patterns we identified in the burden of emergency and operative conditions over time.

### 4.3 Limitations

To our knowledge, this is the first study to map the burden of emergency and operative conditions at the subnational level in India; however, it has several limitations. All data from the GBD 2023 study are estimates. Although GBD estimates are rigorously tested and validated, they may still not accurately capture the true numbers of deaths and DALYs for the conditions we were interested in or for our locations of interest. Furthermore, the GBD study grouped all Union Territories, excluding Delhi, into a single location, and grouped Ladakh with Jammu and Kashmir. This limited our ability to compare Union Territory estimates with those of other states. Additionally, the definitions we used to classify emergency and operative conditions were very broad and likely overestimated the burden of these conditions. For example, our definition of operative care included conditions that were often treated surgically but could also be treated non-surgically. We used the definitions provided by Wimmer et al. because it was the most recent study on the burden of emergency and operative conditions, but a similar analysis using different definitions or frameworks could strengthen our findings. In general, our broad inclusion criteria mean our estimates could be viewed as “worst-case scenario” burden estimates for the emergency and operative conditions. In other words, the burdens should not exceed our estimates. We did not analyze the effects of the COVID-19 pandemic on the burden of emergency and operative conditions, but we know the pandemic caused a brief disruption of healthcare systems worldwide, including challenges to both emergency and operative care. However, the modeling system used in the GBD 2021 study accounts for the effects of the COVID-19 pandemic on mortality and DALYs, and studies have shown that other macro trends have continued to play a significant role in health systems despite the pandemic [20]. We also did not include a cause-by-cause sensitivity analysis, as it was outside the scope of this study. Finally, although we were interested in emergency, critical, and operative (ECO) care as a whole, we did not estimate the burden of critical conditions. The GBD lacked specific data on several of the conditions, such as sepsis and mechanically ventilated patients, that are generally accepted as critical illness syndromes. Thus, we could not accurately capture the burden of critical conditions and therefore excluded it from this analysis. Finally, because we lacked posterior draws and exact covariance structures, we reconstructed distributions from summary statistics. These approximate, but do not reproduce, the full GBD posterior and its covariance. Further, the age- and sex-specific results are limited to India in 2023 and are presented as a reconciled national decomposition rather than a subnationally resolved analysis. Even so, our uncertainty analysis ensured that all estimated values are internally consistent.

## 5 Conclusion

Emergency and operative conditions contribute significantly to India’s overall burden of disease. Although the national morbidity and DALY rates for emergency and operative conditions in India have decreased over time, some states have experienced persistently high burden and/or stagnation in their morbidity and mortality rates. These findings have important implications for healthcare policy in India. The high burden of emergency and operative conditions is most likely a downstream effect of poor emergency and operative care, underscoring the importance of these services in global health.

## Data Availability

All data produced are available online at https://doi.org/10.5281/zenodo.21907392

## 7 Declarations

### Author Contributions

Riley Gerardo: Formal Analysis, Data Curation, Writing – Original Draft, Writing – Review and Editing

Siddhesh Zadey: Conceptualization, Methodology, Writing – Review and Editing, Supervision, Project Administration

Ritika Shetty: Writing – Original Draft, Writing – Review and Editing

Anoushka Arora: Writing – Original Draft, Writing – Review and Editing

Uma Gupta: Writing – Review and Editing

Shirish Rao: Writing – Review and Editing

Aiman Afsar: Writing – Review and Editing

Catherine A Staton: Writing – Review and Editing

Joao Ricardo Nickenig Vissoci: Writing – Review and Editing, Supervision

## Acknowledgements

None

## Funding

None

## Competing Interests

Siddhesh Zadey is the co-founding director of the Association for Socially Applicable Research (ASAR). He also represents ASAR at the G4 Alliance Permanent Council. He serves as the Chair of the Asia Working Group, the G4 Alliance, Fellow of the Lancet Commission on a Citizen-Centered Health System for India, and the Drafting Committee Member for Maharashtra State Mental Health Policy. Other authors declare no competing interests.

## Ethics approval and consent to participate

This manuscript does not involve any human/animal subjects for research. We use publicly available aggregate data. Hence, ethics approval and consent obligations are not applicable.

## Consent for publication

Not applicable

## Availability of data and materials

Data used and produced in the manuscript are available at the Zenodo repository: https://doi.org/10.5281/zenodo.21907392

## AI use declaration

The authors did not use generative AI to draft or edit the manuscript. Grammarly was used for proofreading. We used Claude Cowork Opus 4.8 for statistical simulations and code validation.

## Notes

### Summary of Updates

We have updated the uncertainty analysis, added age- and sex-stratified national estimates for 2023, and removed the sensitivity analysis. Overall, findings remain qualitatively unchanged from the last iteration. However, the estimates now have 95% uncertainty interval bounds.

